# Exploring the Interplay of Poverty, Behavioral Factors, and Obesity: Implications for Diabetes Prevalence in the United States

**DOI:** 10.1101/2025.01.22.25320977

**Authors:** Swarn Chatterjee

## Abstract

The prevalence of diabetes and obesity are a significant public health concern in the United States, with profound implications for millions of Americans. As of 2018, around 34.2 million individuals were diagnosed with diabetes, while an additional 88 million adults were classified as prediabetic, indicating a troubling trend exacerbated by rising obesity rates (Francis, 2021; Liu et al., 2023). Behavioral factors, including smoking, and heavy alcohol consumption are also associated with the prevalence of diabetes and obesity. This study uses geo-spatial county-level data obtained from merging the 2021 CDC PLACES dataset and the 2019 U.S. Census Bureau’s American Community Survey dataset to analyze the roles of smoking, poverty, and binge drinking on the prevalence of obesity and diabetes across the United States. The findings indicate that higher rates of poverty, smoking and alcohol consumption maybe associated with higher rates of obesity, and diversity at the county level. But, the findings of this study also suggest substantial regional variations in these associations.

## Introduction

The prevalence of diabetes and obesity is disproportionately higher among low-income populations and specific racial and ethnic groups, revealing stark socioeconomic disparities in health outcomes (Kim et al., 2023). Research indicates a dramatic increase in diabetes prevalence among individuals living in poverty, with a 100.4% rise noted from 2011 to 2014, further underscoring the critical role of socioeconomic status (SES) in shaping health risks (Chen, 2021). Past research also shows that smoking and alcohol use patterns reveal varying impacts across different demographic groups (Kim et al., 2023; Traversy & Chaput, 2015). Therefore, addressing the complex interplay between socioeconomic factors, behavioral choices, and systemic health inequities is paramount for effective intervention strategies aimed at curbing the obesity and diabetes epidemic in the United States.

Geographic disparities also contribute to the problem, with rural areas facing heightened challenges related to poverty, food insecurity, and healthcare access compared to urban populations (Caldwell et al., 2016; Chang et al., 2022). The ongoing COVID-19 pandemic has further exposed the vulnerabilities of individuals with obesity, as it is associated with heightened severity of COVID-19 infections, emphasizing the need for targeted public health interventions to mitigate these risks (Francis, 2021). Public health policies promoting equitable access to health resources and healthier lifestyle options, particularly in communities most affected by these chronic conditions are essential for building resilient communities (Chang et al., 2022; Chen, 2021; Francis, 2021; Liu et al., 2023).

### Prevalence of Obesity and Diabetes

Diabetes is a chronic health condition that affects the body’s ability to convert food to energy. Typically, the food consumed is broken down into sugar, which is then converted into energy through insulin released in the pancreas. For those suffering from diabetes this process of insulin release or the conversion of sugar to energy malfunctions resulting in a number of other diseases such as heart problems, kidney malfunction, vision impairment etc. (Paulo et al., 2021). The prevalence of diabetes and obesity in the United States has reached alarming levels, significantly affecting public health. As of 2018, approximately 34.2 million Americans were diagnosed with diabetes, and an additional 88 million adults were classified as prediabetic (Liu et al., 2023).

Obesity is defined as a condition where the accumulation of excess body fat can adversely affect the health and well-being of an individual (Kopelman, 2000). Obesity is a primary risk factor for type 2 diabetes, with its effects exacerbated by a sedentary lifestyle. Excess body weight increases insulin resistance in muscle and tissue cells, creating a cycle that further complicates the management of blood glucose levels (Joseph & Ma, 2024). Research indicates that obesity plays a crucial role in the development of Type 2 diabetes, accounting for 30-53% of new cases annually (Francis, 2021). The link between these two conditions has been particularly pronounced over the last two decades, highlighting obesity as a major driver of diabetes incidence (Francis, 2021; Klein et al., 2022).

### Socioeconomic Disparities

The distribution of diabetes and obesity is not uniform across the population; rather, it exhibits substantial disparities based on socioeconomic status (SES) and race/ethnicity. Studies have shown that lower-income individuals are disproportionately affected, with a 100.4% increase in diabetes prevalence among those classified as poor from 2011 to 2014 compared to their high-income counterparts (Kim et al., 2023). These disparities are compounded by factors such as access to traditional financial services, healthcare, and healthy food options, which are often limited in low-income neighborhoods (Chatterjee & Fan, 2023; Kim et al., 2023; Liu et al., 2023). Moreover, obesity-related diabetes incidence varies significantly by gender and racial/ethnic backgrounds, with non-Hispanic white females experiencing the greatest impact (Cameron et al., 2021; Francis, 2021).

Past studies have shown that people living in communities with higher rates of obesity are more likely to be obese due to various neighborhood related factors that maybe mediating the risk of developing obesity among individuals living in areas of higher prevalence of obesity (Ludwig, et al., 2011). In previous literature, obesity has been associated with social and behavioral factors such as poverty, smoking, and problem drinking behavior (Tekin et al., 2018; Strine et al., 2008).

### Behavioral Factors

Behavioral factors also contribute to the prevalence of diabetes and obesity. Evidence suggests that health-related behaviors, such as physical activity, smoking, and dietary choices, play a critical role in determining individual risk levels (Kim et al., 2023; Liu et al., 2023). A higher intake of ultra-processed foods has been associated with an increased risk of Type 2 diabetes, indicating that dietary patterns are significant in the discussion of these health issues (Duan et al., 2022). Additionally, smoking and alcohol consumption patterns differ across socioeconomic and racial/ethnic groups, further influencing health outcomes (Kim et al., 2023; Tekin et al., 2018).

Alcohol consumption and smoking are additional lifestyle factors that interact with diabetes risk. Alcohol use has been identified as a significant mediator in the SES-diabetes relationship, particularly among males, where higher consumption is linked to greater risk (Liu et al., 2023). In middle-aged populations, smoking rates are affected by social pressures, with lower SES individuals facing greater challenges in cessation efforts (Liu et al., 2023). Similarly, in past literature, obesity has been associated with social and behavioral factors such as poverty, smoking, and problem drinking behavior (Tekin et al., 2018; Strine et al., 2008).

## Methodology

### Data

This study used the ESRI ArcGIS to analyze county-level data on obesity, poverty, binge drinking, and smoking in the United States, and their relationship with diabetes prevalence. The methodology was designed to ensure accurate geospatial representation, data integration, and analysis to identify patterns. The county-level data for obesity prevalence, poverty rates, binge drinking rates, smoking rates, and diabetes prevalence were obtained from the Centers for Disease Control and Prevention (CDC) PLACES dataset (CDC, 2021). The income and poverty data was obtained from the American Community Survey (U.S. Census, 2019).

### Data Integration and Mapping

The data were imported into ArcGIS, and geocoding was performed to ensure the data were correctly mapped to the corresponding county codes. Thematic maps were then created for each variable (obesity, poverty, binge drinking, smoking, and diabetes).

### Data Visualization and Interpretation

Multi-layer maps were then created by overlaying diabetes prevalence with obesity, poverty, binge drinking, and smoking rates to visually identify spatial patterns and potential relationships. Additional multi-layer maps were also created to examine prevalence of smoking and obesity, smoking and diabetes, binge drinking and obesity, binge drinking and diabetes, income and obesity, income and diabetes.

This methodology leveraged ArcGIS’s powerful spatial visualization capabilities to examine the complex relationships between diabetes and various risk factors at the county level. By integrating multiple data sources and employing spatial visualization techniques, valuable insights were gained by exploring the geographic distribution of diabetes and its associated risk factors across the United States.

## Results

Obesity rates by county are displayed in figure 1. The results indicate that higher rates of obesity are concentrated around Southeastern United States. However, areas of the upper-Midwest, and across the East coast, as well as pockets of the West coast also have higher rates of obesity. This indicates that obesity is indeed a nation-wide problem, and a serious public health challenge that risks the overall health and well-being of the U.S. population.

**Figure 1:**
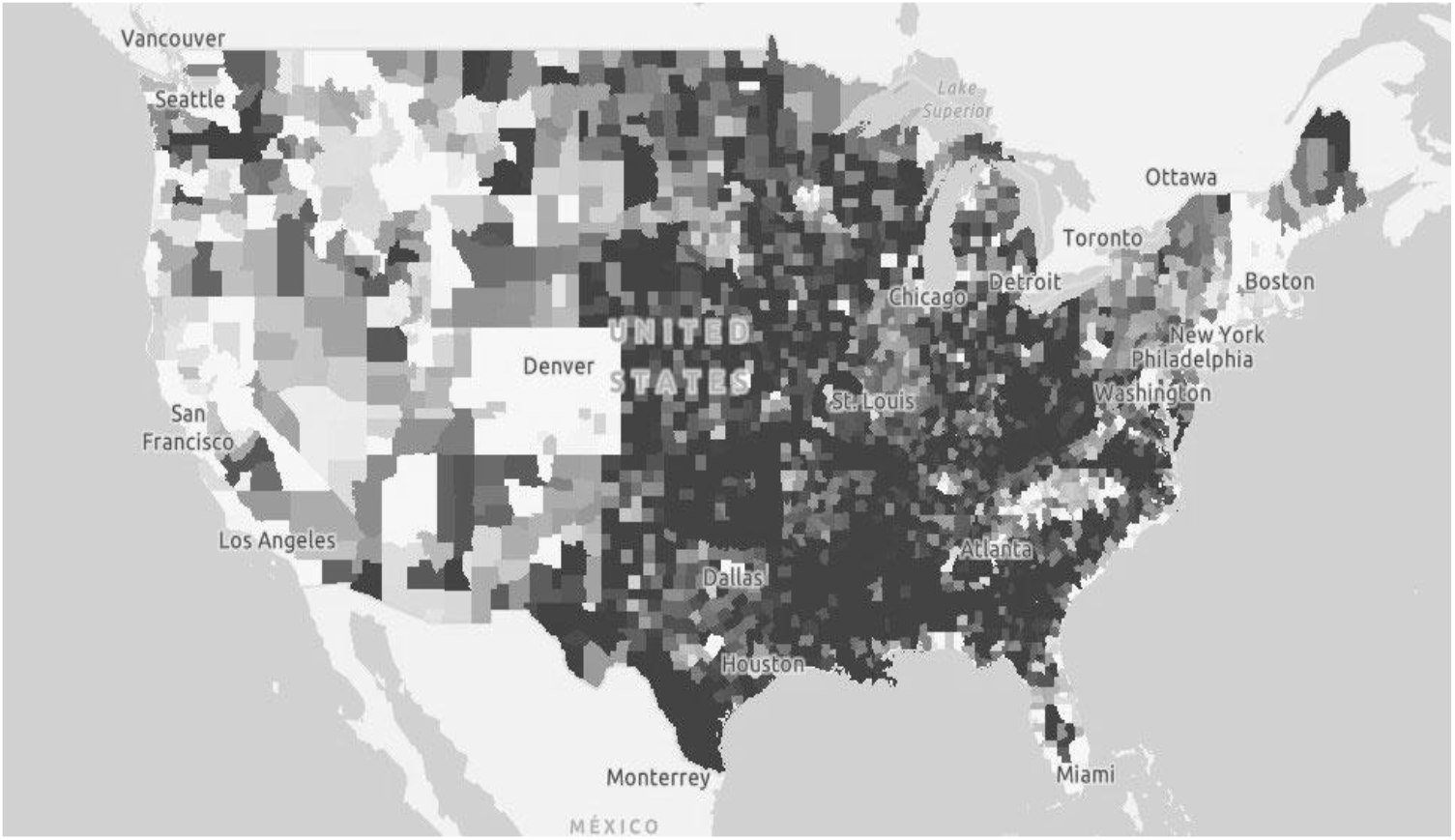
Obesity Rates by County

Figure 2 shows the diabetes rates by county. As can be seen from the figure, diabetes is prevalent in areas of the Southeastern United States where obesity is also a concern. Interestingly, the areas of upper Midwest where obesity was also prevalent, does not have the same high rate of prevalence of diabetes. Diabetes is also prevalent in areas of the Pacific Northwest and the West coast where obesity was not prevalent. These findings suggest that while obesity might be a factor in the prevalence of diabetes, there are possibly other correlates of the occurrence of diabetes in places where obesity rates might be low.

**Figure 2:**
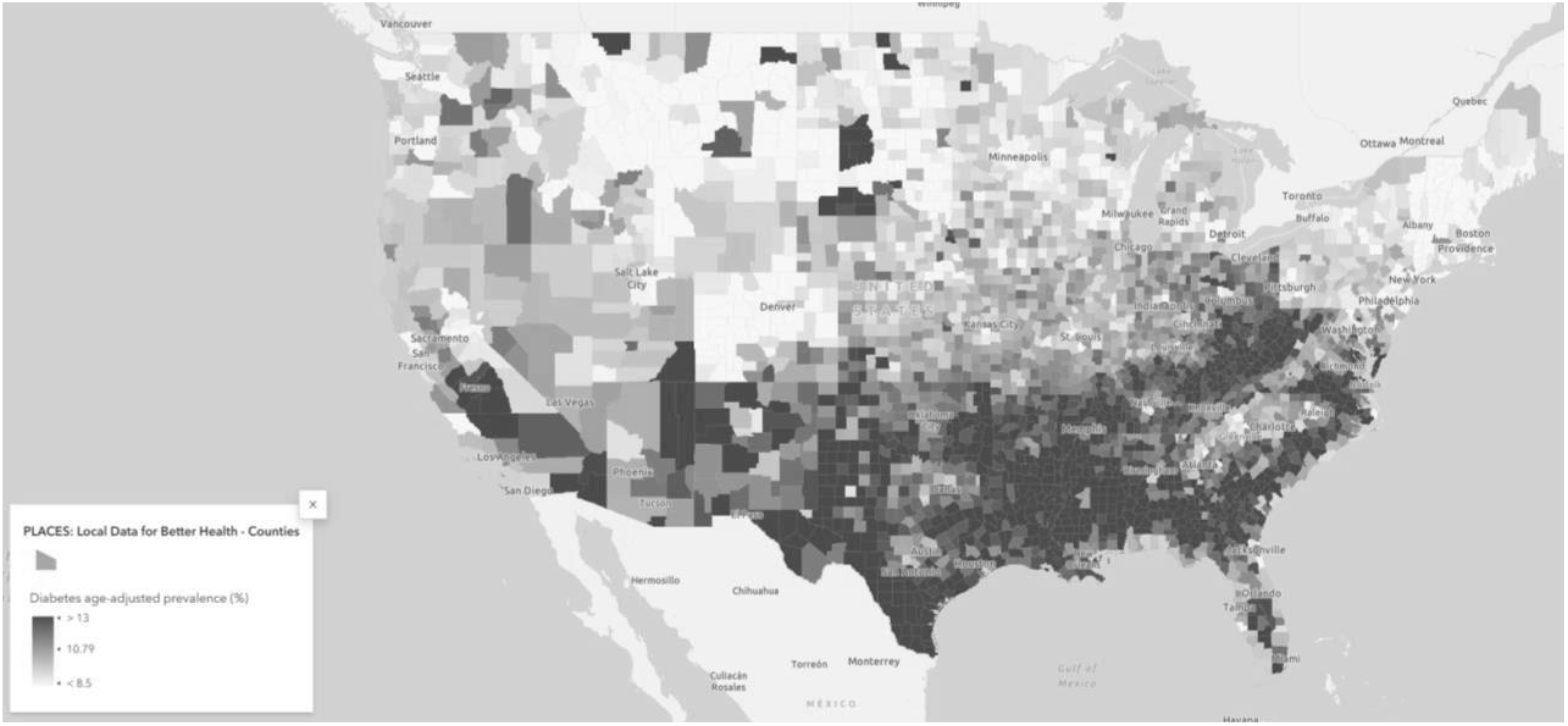
Diabetes Rates by County

Figure 3 shows the prevalence of smoking by county. It can be seen from the figure that the rate of smoking is higher in the Eastern half of the United States. The percentage of smokers tend to be lower on the West coast, especially California, Utah, and Colorado for example. In past literature, obesity has been associated with social and behavioral factors such as poverty, and smoking (Tekin et al., 2018; Strine et al., 2008).

**Figure 3:**
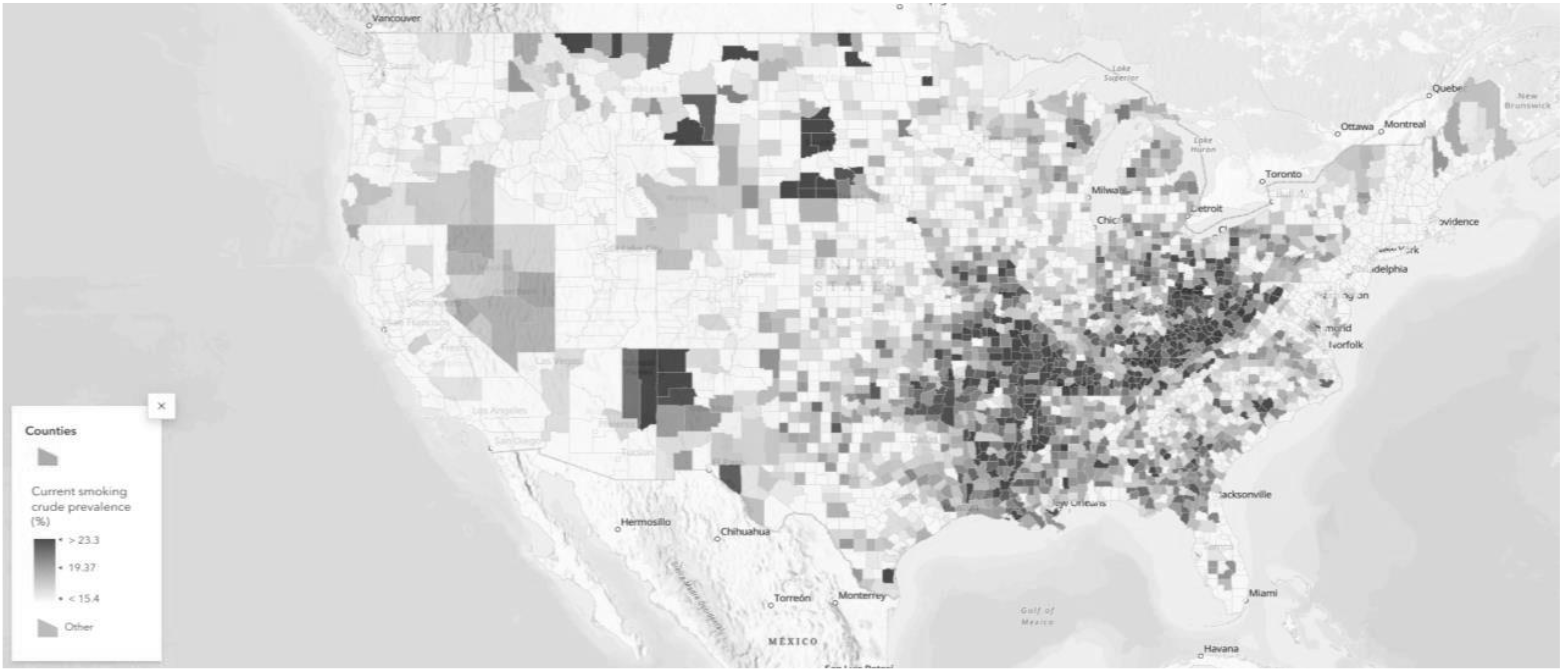
Prevalence of Smoking by County

A multi-layer map of obesity rates and smoking is shows in figure 4. The larger circles indicate greater age adjusted smoking prevalence among adults. The results show that the areas with higher rates of obesity also correlate with areas where smoking is prevalent among a larger section of the population. The Southeast, Midwest, and parts of the East coast have a higher concentration of areas with very high rates of obesity and a higher rate of smoking prevalence in the population.

**Figure 4:**
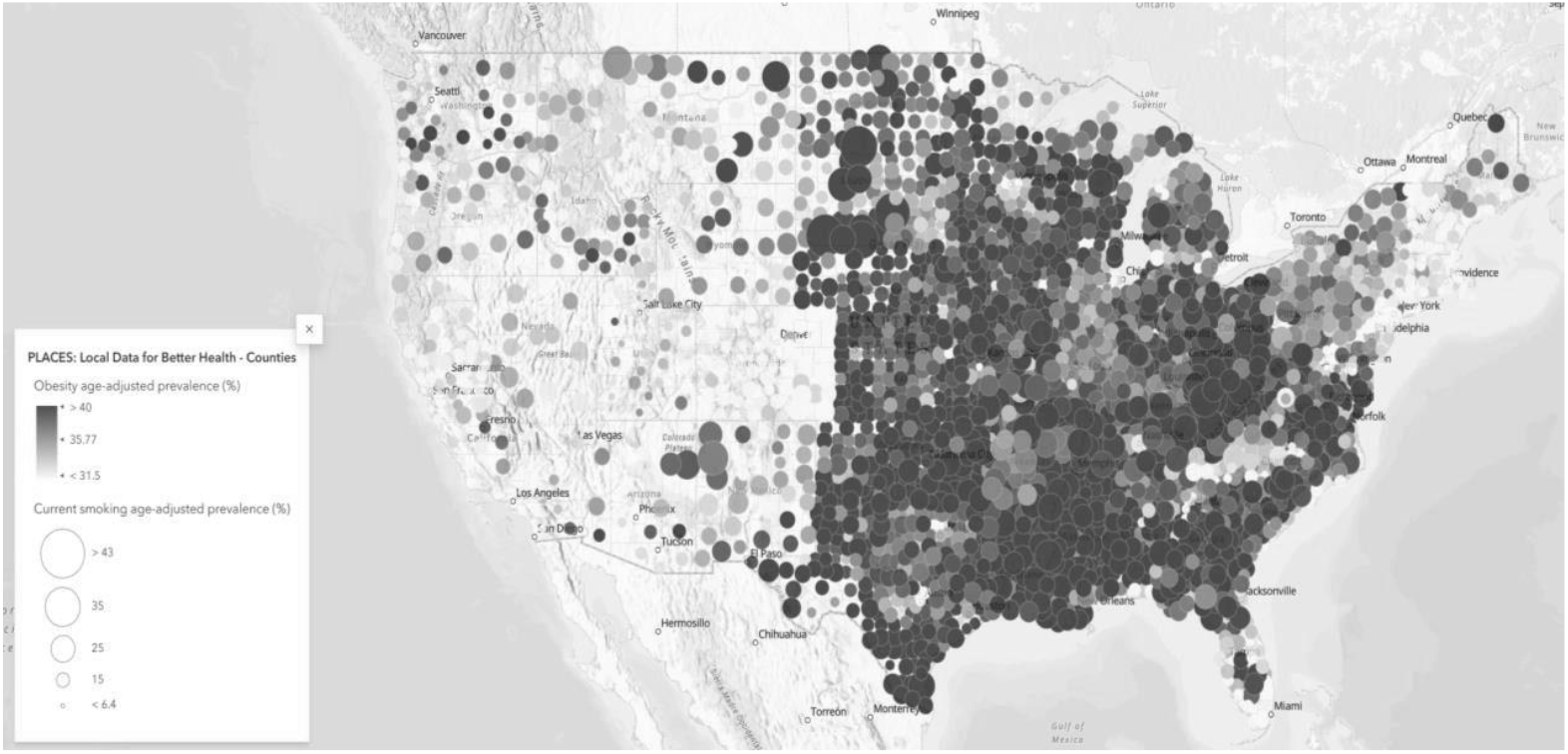
Obesity and Smoking

Another multi-layer map overlaying smoking and diabetes is constructed in figure 5. The results indicate that although there is a visible overlap between smoking and diabetes in the Southeastern and Eastern United States, the results appear mixed in the upper midwestern and western United States. Utah, which has low rates of smoking and low prevalence of obesity, appears to have a higher prevalence of diabetes in some of the counties.

**Figure 5:**
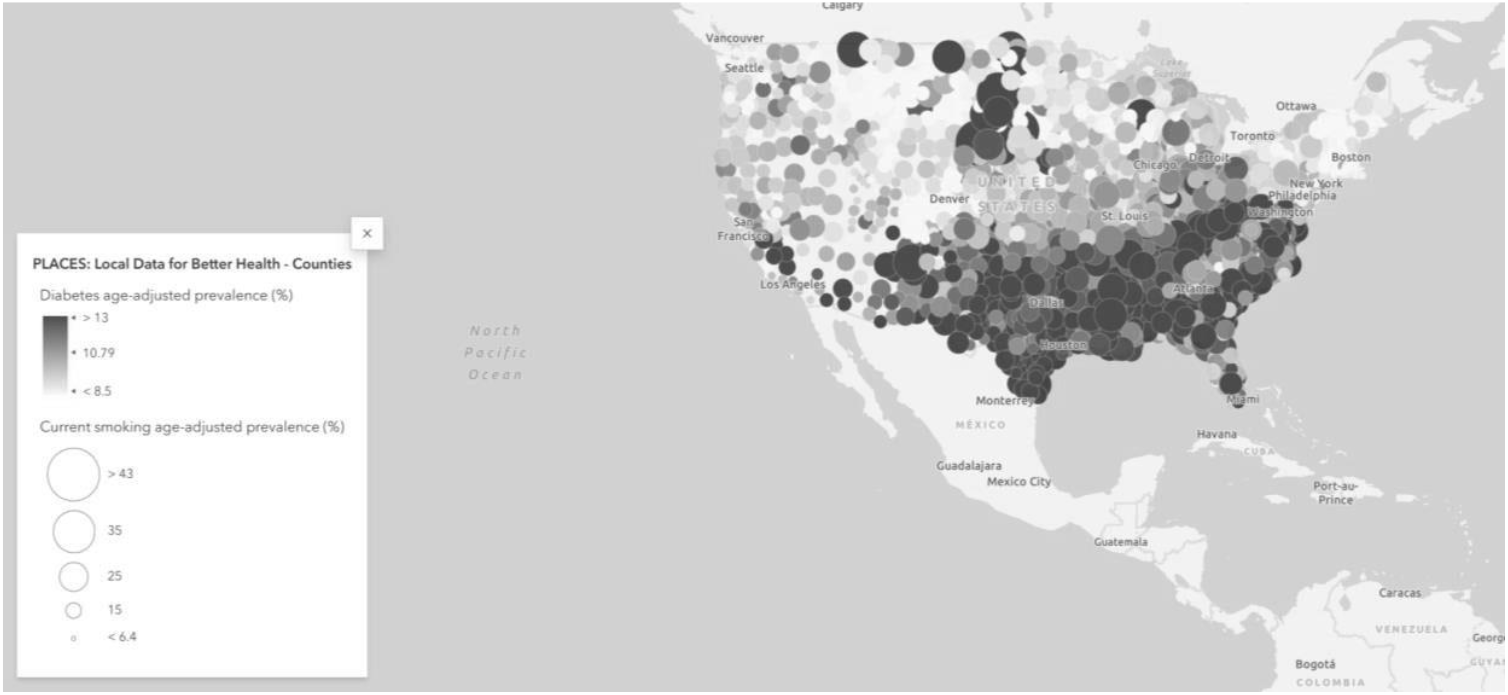
Diabetes and Smoking

Figure 6 presents the map for binge drinking by county. The results indicate that binge drinking is more prevalent in the Western and Mid-western sections of the United States. Another association that is observed is that binge drinking appears concentrated around urban areas. The Southeastern United States, where the residents are considered to be more socially conservative, have higher rates of smoking (figure 4) but lower rates of heavy drinking (figure 5). Utah stands out as the state having amongst the lowest rates of heaving drinking or smoking.

**Figure 6:**
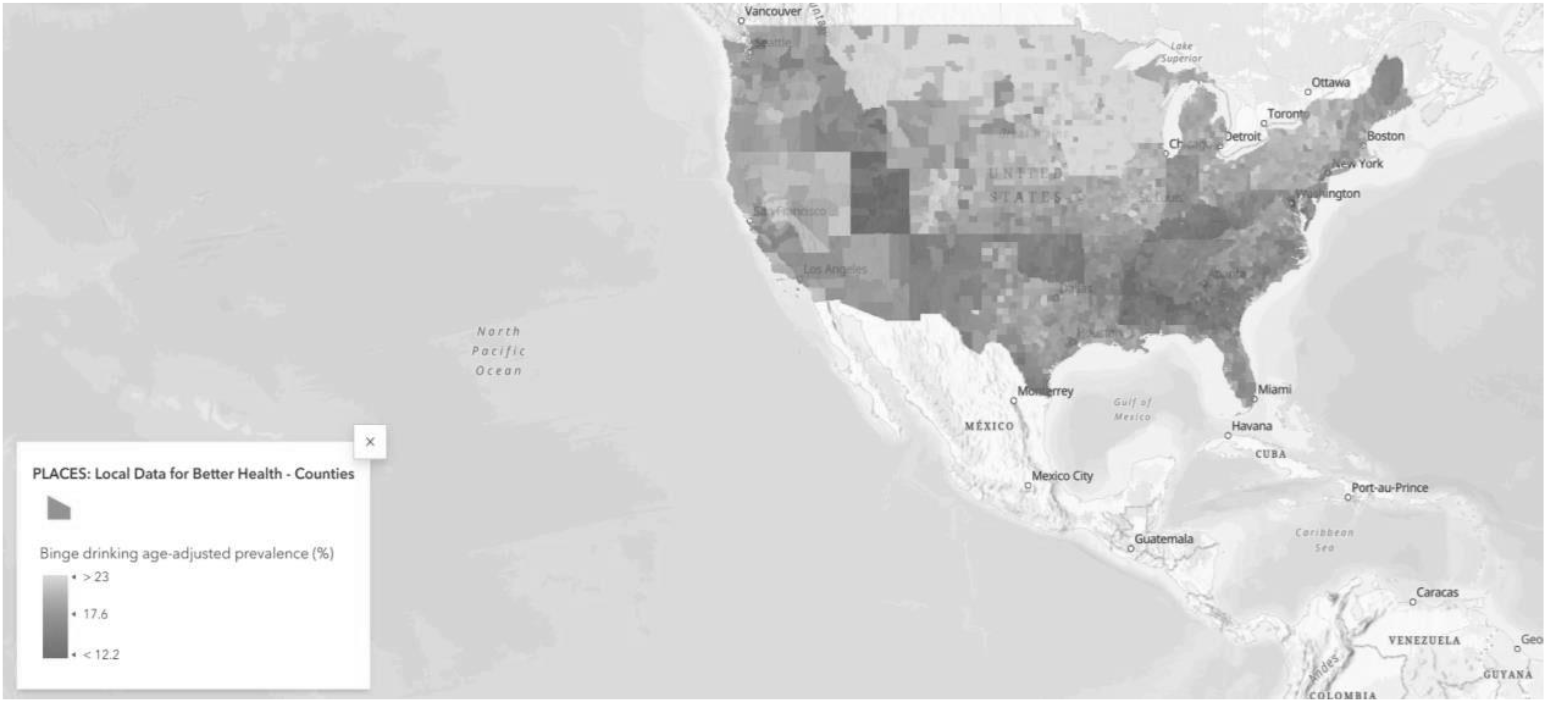
Binge Drinking by County

Figure 7 shows an interesting relationship between binge or problem drinking and obesity. Although, there appears to be a correlation between binge drinking and obesity in the western and upper midwestern United States, the association does not seem as obvious in the south. Although many areas of the southeastern United States had high rates of obesity, the prevalence of binge drinking in the south was relatively low.

**Figure 7:**
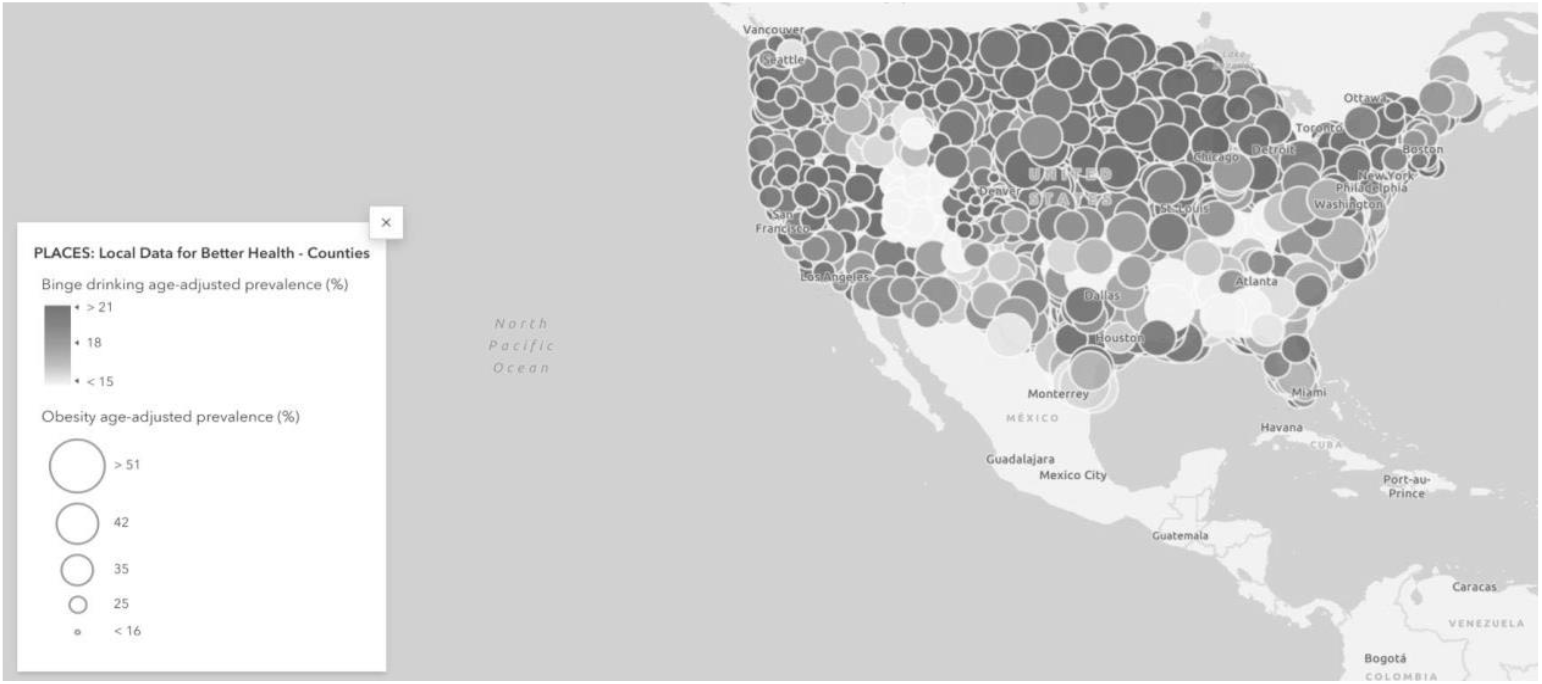
Binge Drinking and Obesity

A similar association is also observed when examining diabetes and binge drinking. As we can see from figure 8, although the midwestern United States has a higher prevalence of binge drinking, the association with the prevalence of diabetes is not as obvious. The southeastern United States on the other hand does have high rates of diabetes across many counties, but does not have a high prevalence of binge drinking.

**Figure 8:**
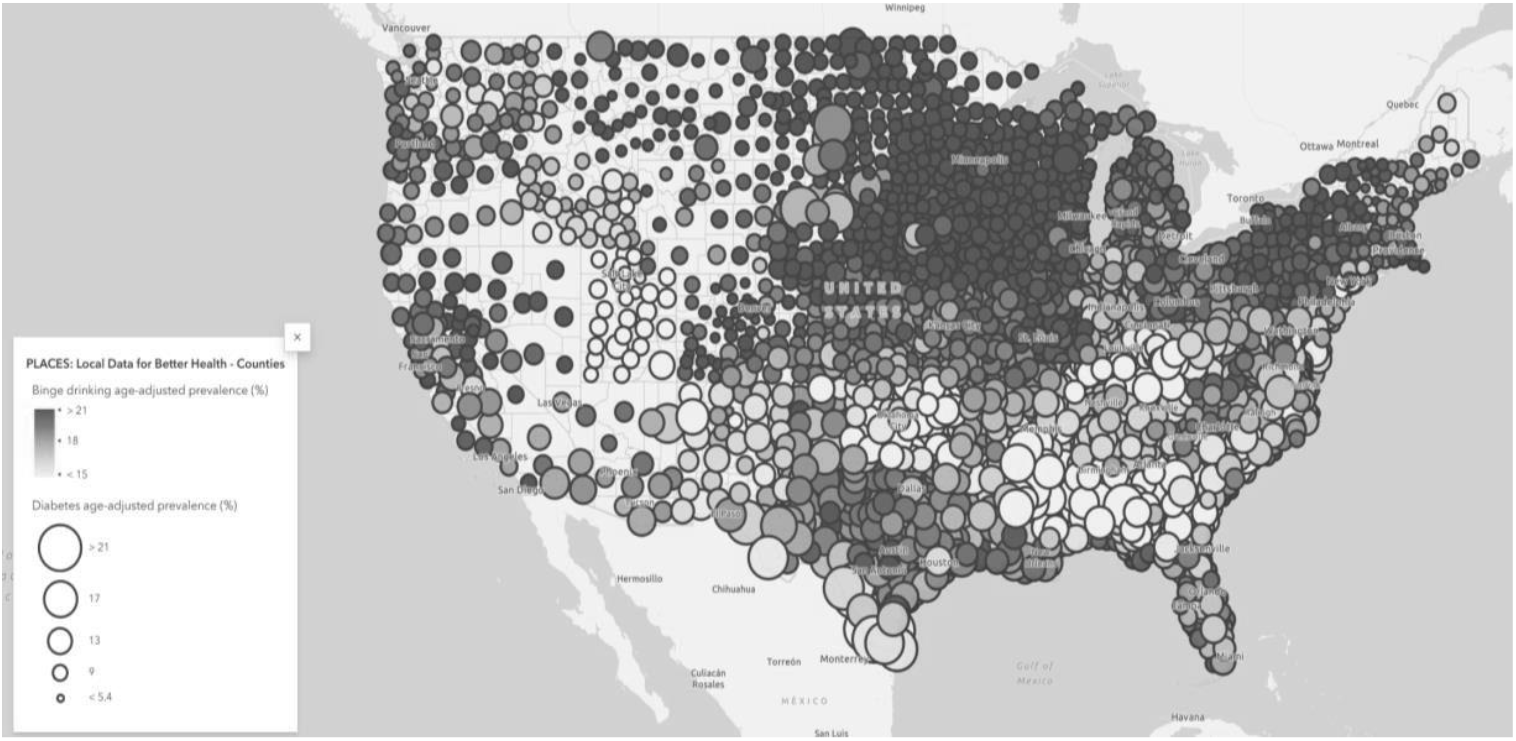
Binge Drinking and Diabetes

Figure 9 shows the percent of county-level population whose income in the past 12 months is below poverty level. As indicated by the map legend, we can see that the counties marked in darker shades have more poverty.

**Figure 9:**
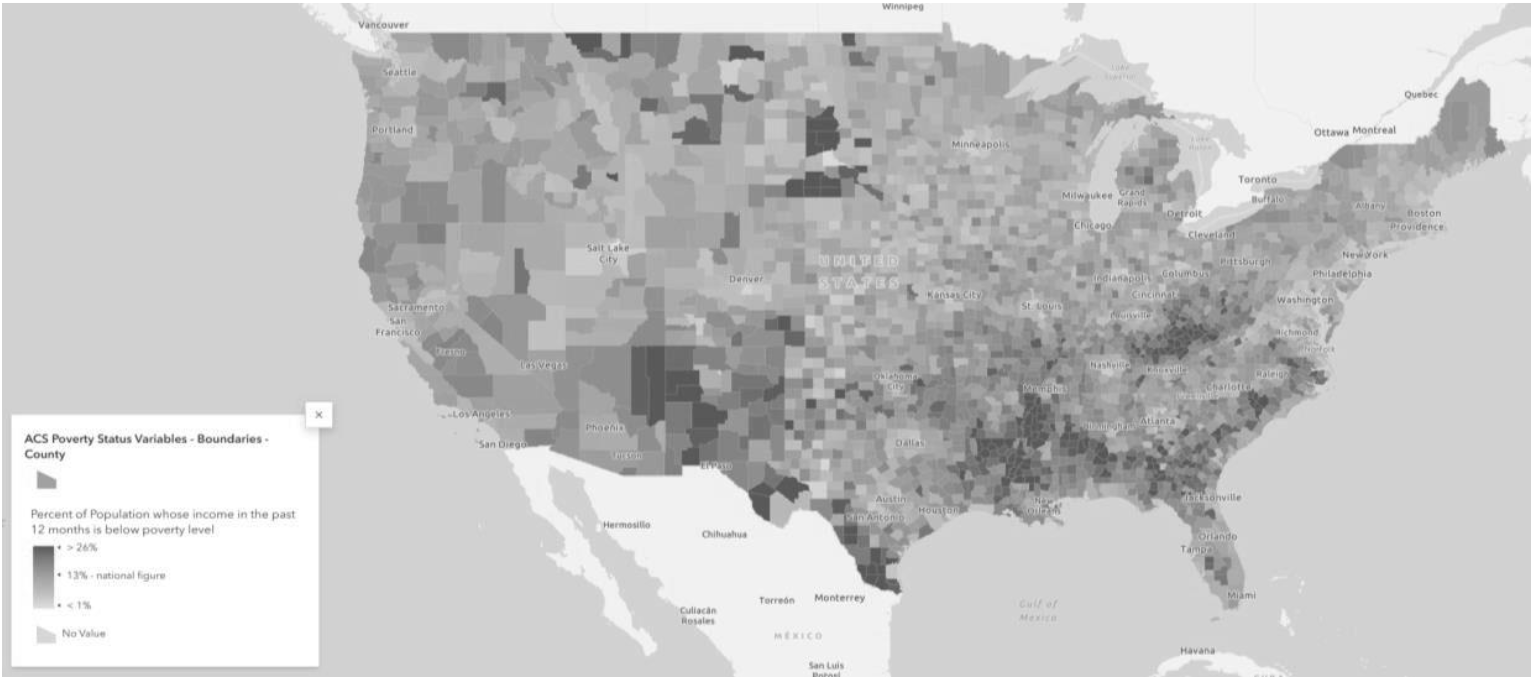
Percent of Population Living in Poverty

Counties with higher rates of poverty appear frequently in the Southern and the mountain west regions of the United States. The next map shows the relationship between income and obesity. Figure 10 shows a multi-layer map of income and obesity.

**Figure 10:**
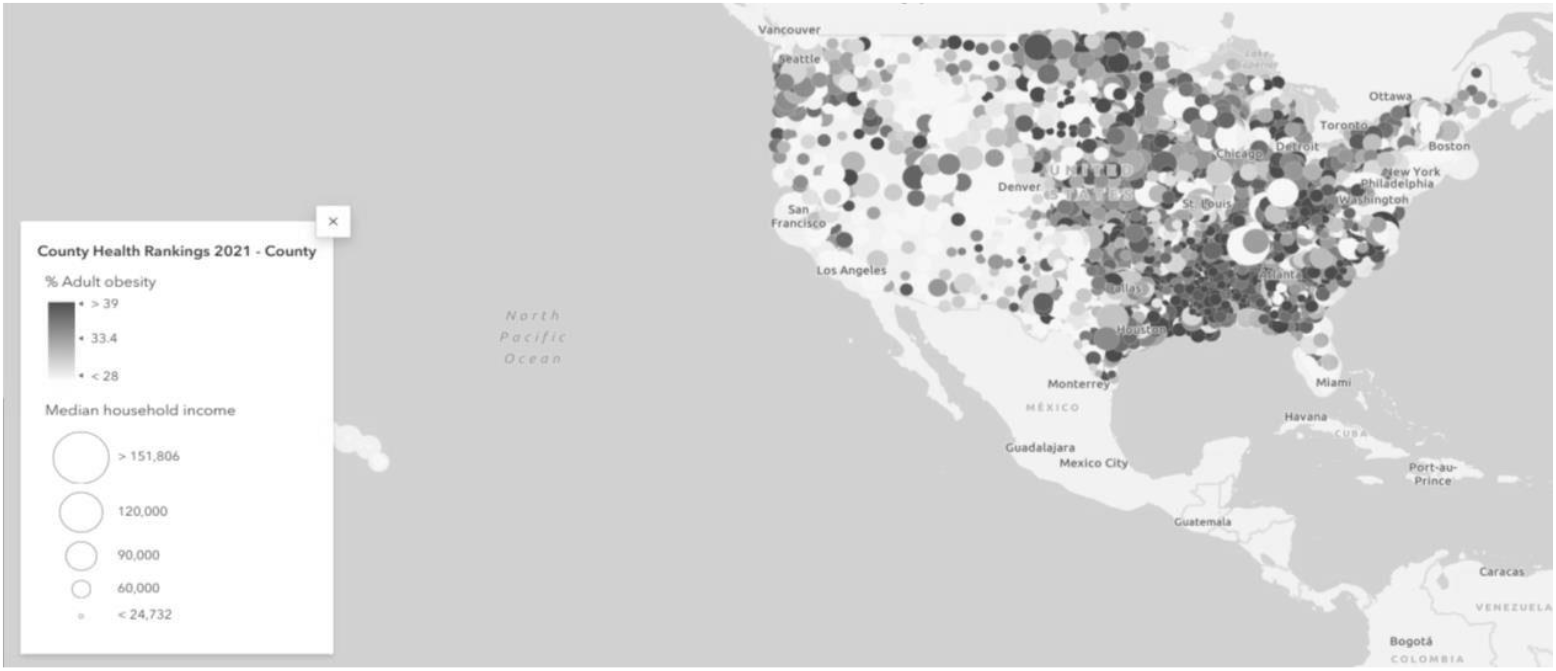
Income and Obesity

The results from figure 10 indicate that the counties with higher rates of obesity also tend to have lower levels of median household income. The association is visibly strong in areas of the Southeastern United States. On the contrary, in some areas of the upper Midwest and the Pacific northwest, obesity appears prevalent even among individuals with higher median income.

The map shown in figure 11 indicates an association between diabetes and income. Higher levels of diabetes are associated with lower levels of median household income. And the map in figure 12 shows the association between obesity and prevalence of diabetes.

**Figure 11:**
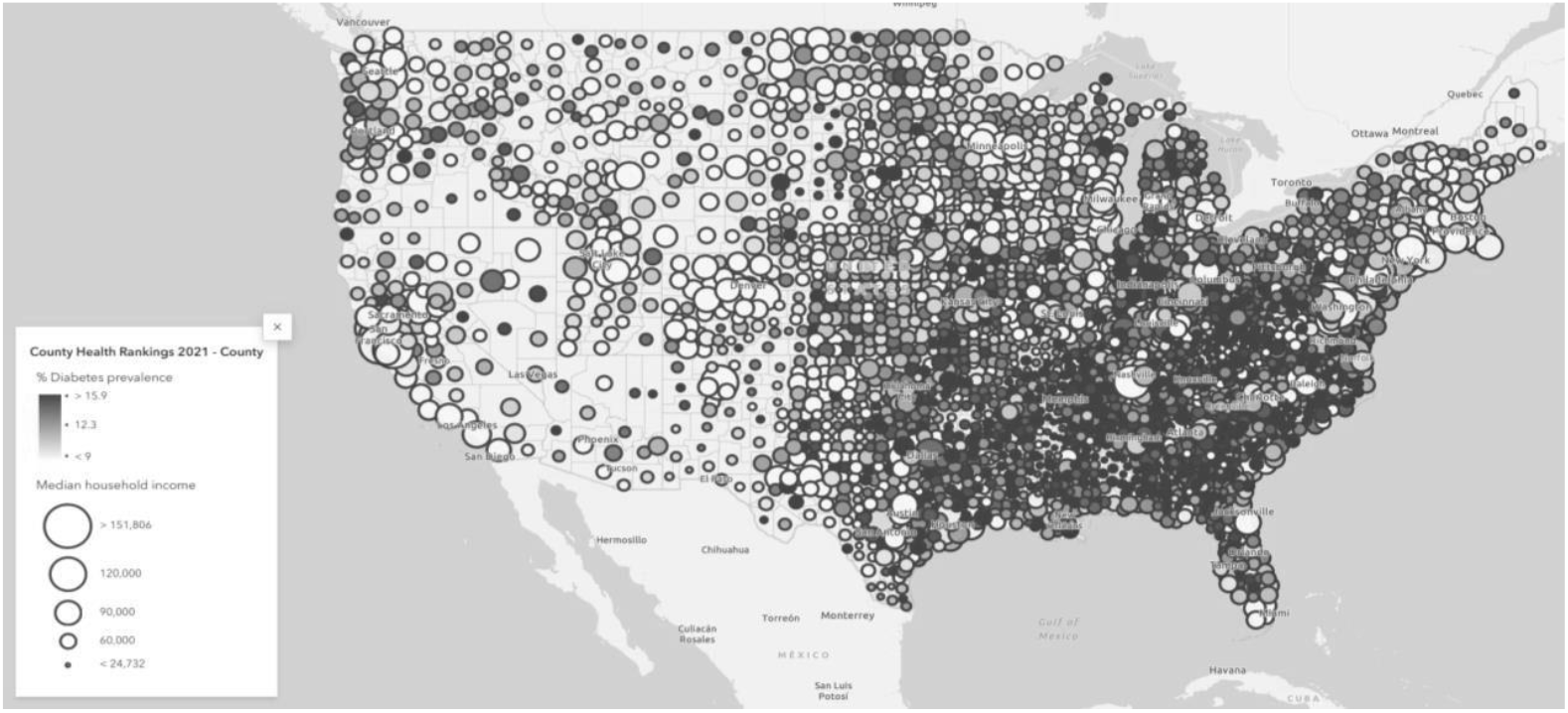
Income and Diabetes

**Figure 12:**
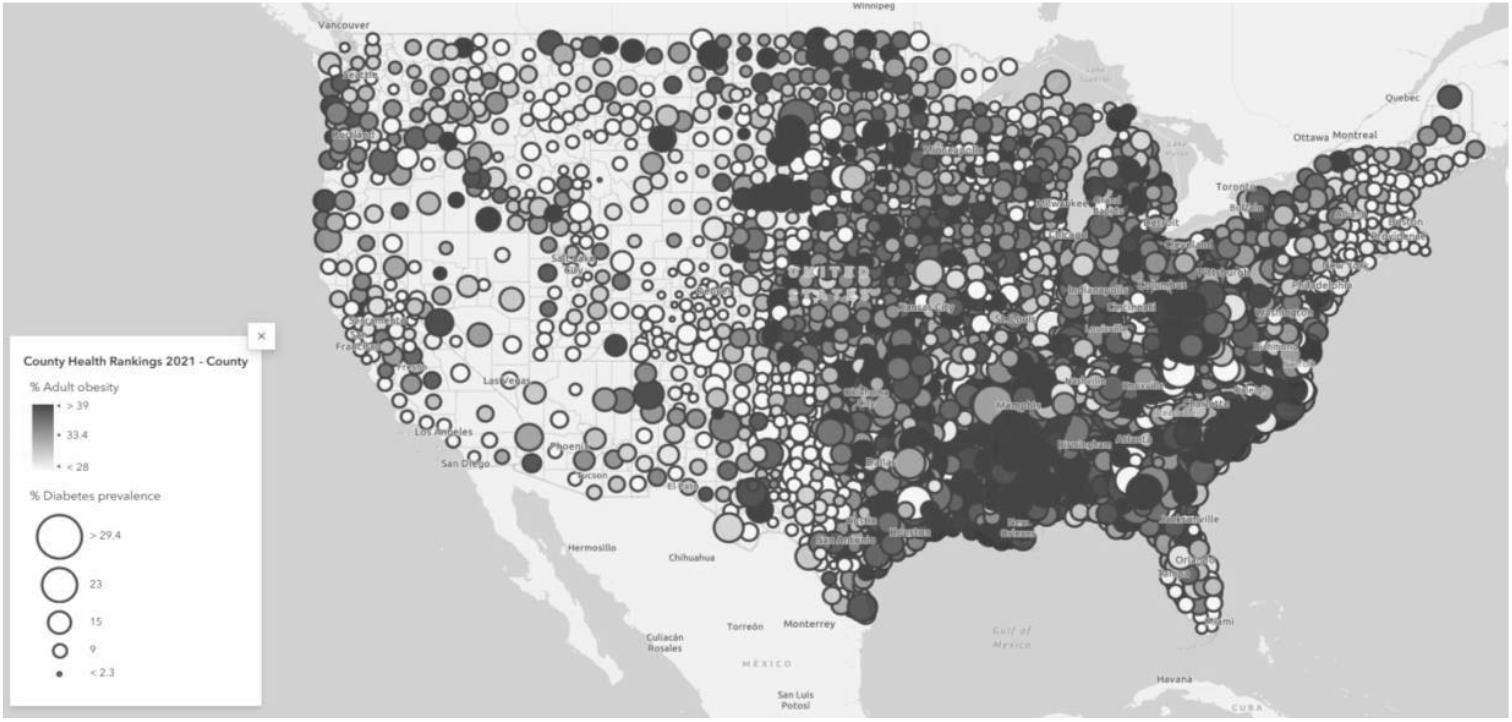
Obesity and Diabetes

In this map, there appears to be a very visible linkage between obesity and diabetes across the country. The residents of the Southeastern and Midwestern United States face the twin risks of both diabetes and obesity more than any other region of the United States.

## Conclusion

In conclusion, the findings of this study indicate a substantial overlap between counties with high rates of obesity and diabetes, and counties with higher rates of poverty, smoking, and alcohol consumption. These findings underscore the complex interplay between health behaviors, socioeconomic factors, and geographic disparities (Caldwell et al., 2016; Liu et al., 2023; Richards et al., 2022). While risky health behaviors such as binge drinking and smoking are linked to higher rates of obesity and diabetes, income constraints also play a crucial role in shaping these health outcomes (Choung et al., 2022; Chatterjee & Kim, 2023; Worthy et al., 2010). The findings from this study corroborate with findings from past literature that found neighborhoods characterized by high poverty to be associated with higher rates of problematic health behaviors (Richards et al., 2022).

Individuals experiencing poverty may turn to smoking as a coping mechanism (Peretti-Watel et al., 2009), yet this behavior can exacerbate their financial difficulties due to the costs associated with tobacco products. Addressing these challenges requires a multifaceted approach that includes poverty alleviation programs, targeted community health education, and behavior modification initiatives (Chang et al., 2017; 2019). Specific attention to regions such as the Southeastern United States, where obesity and diabetes are both prevalent, could be key to reducing these health disparities. Future research should explore additional factors, including behavioral, attitudinal, and socioeconomic influences, to provide a more comprehensive understanding of the drivers of obesity and diabetes. Ultimately, empowering communities through education and outreach programs could pave the way for healthier behaviors and better long-term health outcomes across the United States.

## Data Availability

All data produced are available online at
https://www.cdc.gov/places/methodology/index.html
https://www.census.gov/programs-surveys/acs/about.html

https://www.cdc.gov/places/methodology/index.html

## Notes

### Competing Interest Statement

The authors have declared no competing interest.

### Funding Statement

This study did not receive any funding

